# Mental health issues among medical students: Exploring predictors of mental health in Dhaka during COVID-19 pandemic

**DOI:** 10.1101/2023.06.04.23290948

**Authors:** Subigya Man Lama, Md. Toufiq Elahi Ahad

**Affiliations:** Department of Public Health, Faculty of Arts and Social Sciences, American International University – Bangladesh, Dhaka, Bangladesh; Department of Medicine, Dhaka Medical College Hospital, Dhaka, Bangladesh; Maternal and Child Health Division, International Centre for Diarrhoeal Disease Research, Bangladesh, Dhaka, Bangladesh

**Keywords:** Mental health, medical students, COVID-19, Dhaka

## Abstract

**Background:** Mental health has always been under the shadow of everyone’s belief about their health. Concerns about mental health have already risen in the whole world. The COVID-19 pandemic has caused havoc worldwide, notably in the educational system. It has been difficult to quantify the influence of COVID-19 on the mental health of medical students in Bangladesh.

**Aims:** This study was conducted to assess medical students’ mental health status in Dhaka during COVID-19 pandemic.

**Methods:** This study was undertaken at Dhaka Medical College, Dhaka, Bangladesh and 359 medical students were the primary respondents for this study.

**Results:** Depression, anxiety and stress were found in around half of the study participants. Overall, three-fourth of the medical students had poor mental health status. The research study showed that depression, anxiety and stress were dependent on various socio-demographic and behavioral characteristics of medical students.

**Conclusion:** Poor mental health is still highly prevalent in the medical students. Different factors like age, gender, academic year, and physical exercise behavior have affected medical students’ mental health. This calls for attention towards the needs of the more vulnerable demographics and creating a welcoming environment for medical students.

## Introduction

Mental health has always been under the shadow of everyone’s health beliefs. According to World Health Organization, “Health is a state of complete physical, mental and social well-being and not merely the absence of disease or infirmity.” [1] Along with physical health, this concept encompasses mental and social well-being. Concerns about the mental health have already risen in whole world. Depression is one of the greatest reasons of disability in the world [2]. In people aged 15 to 29, suicide is the fourth highest cause of mortality [3].

Before joining M.B.B.S. course, medical students already have to deal with a stressful entrance exam [4]. After joining the M.B.B.S. course, the burdens of various new medical subjects are added along with everyday exams. Medical studies have always been one of the toughest course to study [5].

With the recent COVID-19 pandemic, everything has changed in the world. New methods and new rules were implemented to tackle the pandemic and continue the studies [6]. The most popular study method became virtual learning and it was seen fruitful in learning of medical students [7]. It has been difficult to quantify the influence of COVID-19 on the mental health of medical students in Bangladesh. Medical students are already more prone to poor mental health globally.

Medical students during their course, medical students have high pressure from loads of studies and exams. Repeated everyday exams and exhausting level of study have a delirious effect on medical students’ mental health [8]. With the COVID-19 pandemic, every physical class has stopped and online classes have taken place. Mental health has not been the same as how it has been used to during the physical classes [9,10]. Medical students with poor mental health are vulnerable to dropping out of the medical course [11] and suicidal ideation.

## Materials and Methods

### Design, setting and participants

This was an analytical cross-sectional study conducted at Dhaka Medical College from October till December 2021. Cochran’s formula [12] was used to figure out the sample size for this study, where anticipated prevalence rate of poor mental health was assumed from the findings of the study done by Hasan et al. [13]. Calculating through the formula, 342 samples were enough for the present study. Considering a 5% non-response rate, 359 medical students under M.B.B.S. course were considered and recruited using a convenience sampling approach.

### Measurements

The data were gathered utilizing a structured questionnaire that was handed out to the study participants in person. The questionnaire included socio-demographic and behavioral characteristics, COVID-19 status as well as Depression, Anxiety and Stress Scale – 21 (DASS-21). Any medical students under mild to extremely severe depression, anxiety, or stress were considered having poor mental health.

### Data analysis

The data were examined and double-checked for completeness and correctness. Statistical Package for Social Sciences (SPSS) version 25.0 was used to de-identify, clean, code, categorize, input, and analyze data. To see if there was a link between variables, Pearson chi-squared tests or Fisher’s exact tests (if cell frequency was less than 5) were used. For all tests, the statistical significance threshold was set at p < 0.05 (two-tailed).

### Ethical consideration

The study received ethical approval (Issue Number: 01/2022) from the Ethical Review Committee of American International University - Bangladesh. The study’s background, aims, risks, and benefits were all explained to all participants. Informed written consent were obtained from each of the participants. Only those who were willing to participate and offer informed consent were included in the study. Participants were able to exit the study at any time and were not penalized. The participant’s anonymity and confidentiality were preserved throughout the study.

## Results

Among the 359 participants, age between 21 and 24 years (65.5%) was the majority age group in the study, followed by age less-than and inclusive of 20 years (30.6%). The mean age came in at 21.54 years, with a standard deviation of 1.743. Females made up 58.8% of the study participants, while males made up 41.2%. Most of the study participants were unmarried (94.2%) and having family sizes of two to five members (79.7%). Regarding family income, 21.4%, 46.5%, and 32.0% of the study participants had family income of less than BDT 30,000, between BDT 30,000 & BDT 60,000 and more than BDT 60,000 respectively. Of the total participants, 71.6% resided in hostels, 23.7% resided at home with their parents, and 4.7% resided in flats or apartments. 21.4% of the study participants were in first year, 19.8% were in second year, 18.9% were in third year, 18.7% were in fourth year and 21.2% were in fifth year of their medical studies.

**Table 1:** Characteristics of the study participants (n=359)

| Variable | Frequency | Percentage(%) |
| --- | --- | --- |
| **Socio-demographic characteristics** |  |  |
| Age (years) |  |  |
| ≤ 20 | 110 | 30.6 |
| 21 - 24 | 235 | 65.5 |
| ≥ 25 | 14 | 3.9 |
| Gender |  |  |
| Male | 148 | 41.2 |
| Female | 211 | 58.8 |
| Marital status |  |  |
| Unmarried | 338 | 94.2 |
| Married | 18 | 5.0 |
| Divorced/Separated | 3 | 0.8 |
| Family size |  |  |
| Two to five members | 286 | 79.7 |
| Six to ten members | 71 | 19.8 |
| More than ten members | 2 | 0.6 |
| Family income |  |  |
| Less than BDT 30,000 | 77 | 21.4 |
| Between BDT 30,000 to BDT 60,000 | 167 | 46.5 |
| More than BDT 60,000 | 115 | 32.0 |
| Residence |  |  |
| Hostel | 257 | 71.6 |
| Home with parents | 85 | 23.7 |
| Flat or apartment | 17 | 4.7 |
| Academic year |  |  |
| First year | 77 | 21.4 |
| Second year | 71 | 19.8 |
| Third year | 68 | 18.9 |
| Fourth year | 67 | 18.7 |
| Fifth year | 76 | 21.2 |
| **Behavioral characteristics** |  |  |
| Physical exercise |  |  |
| Yes | 235 | 65.5 |
| No | 124 | 34.5 |
| Hours of sleep |  |  |
| Less than 8 hours | 271 | 75.5 |
| More than 8 hours | 88 | 24.5 |
| Smoking status |  |  |
| Not smoking | 348 | 96.9 |
| Smoking | 11 | 3.1 |
| **COVID-19 related information** |  |  |
| Vaccination status |  |  |
| Unvaccinated | 4 | 1.1 |
| Vaccinated (Single dose) | 5 | 1.4 |
| Vaccinated (Both dose) | 350 | 97.5 |
| Previous COVID-19 infection status |  |  |
| Not infected before | 259 | 72.1 |
| Infected before | 100 | 27.9 |

65.5% of the study participants didn’t have the habit of exercising or playing sports for at least thirty minutes three or more times a week, while 34.5% had the habit. Sleeping hours of less than 8 hours were also seen in 75.5% of the study participants, whereas 24.5% slept longer than 8 hours. Only 3.1% of the study participants had the habit of smoking.

97.5% of the study participants already had both dose of vaccine against COVID-19, whereas 1.4% had single dose of vaccine and 1.1% were still unvaccinated against COVID-19. Of the total participants, 27.9% had been infected with COVID-19 before while 72.1% didn’t have any infection with COVID-19 before.

40.9% of the study participants didn’t have depression, whereas 12.8%, 20.1%, 9.7% and 16.4% had mild, moderate, severe and extremely severe level of depression respectively. 37.3% of the study participants didn’t have anxiety, whereas 9.5%, 25.3%, 11.1% and 16.7% had mild, moderate, severe and extremely severe level of anxiety respectively. 58.8% of the study participants didn’t have stress, whereas 14.2%, 10.6%, 10.9% and 5.6% had mild, moderate, severe and extremely severe level of stress respectively. Overall, 73.8% of the study participants had poor mental health i.e. with either only depression, only anxiety, only stress, or combination.

**Fig 1.**
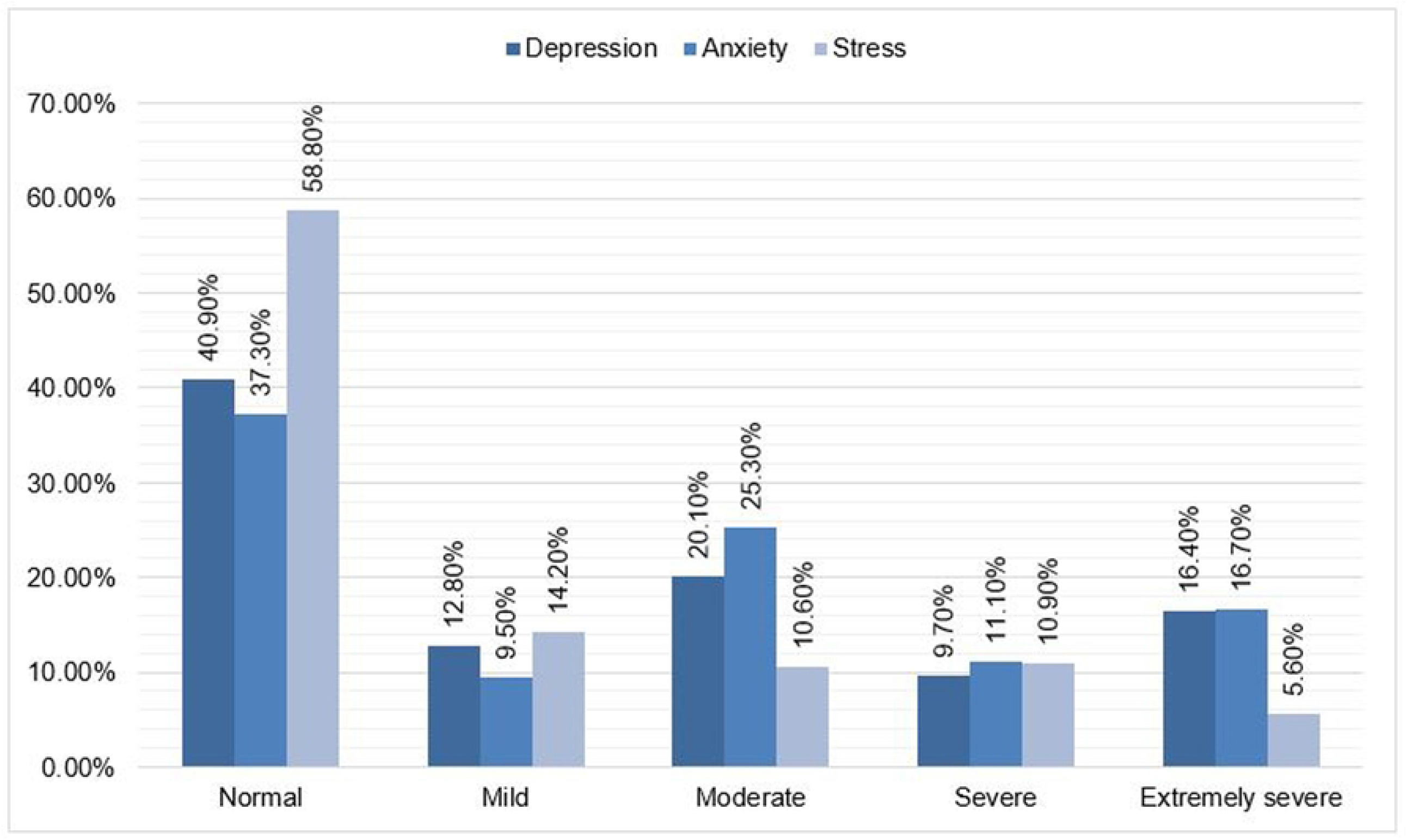
Proportion of severities of depression, anxiety and stress among the study participants (n=359)

**Fig 2.**
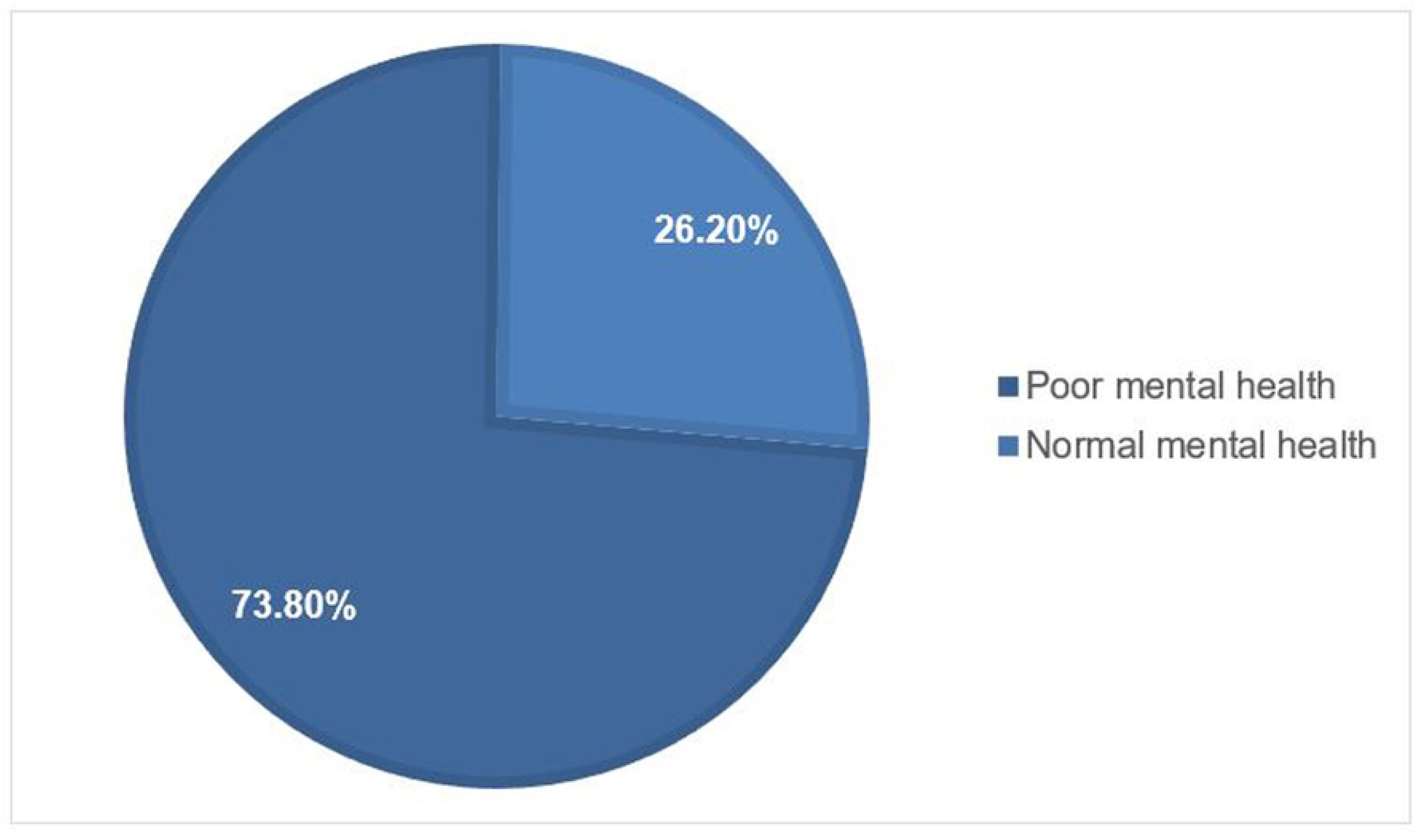
Mental health status of the study participants (n=359)

A higher percentage of individuals aged 21 to 24 years experienced depression (60.8%) compared to other age groups, and this difference was statistically significant (p = 0.034). Although females had a higher rate of depression (62.7%), this was not statistically significant. Unmarried individuals (94.3%), those from families with two to five members (79.2%), and those with a family income between BDT 30,000 and BDT 60,000 (44.3%) were more likely to experience depression, as were those residing in a hostel (68.4%) and in their first year of medical studies (25.0%), although these differences were not statistically significant. Similarly, anxiety was more common in the 21 to 24 age group (59.6%) and was statistically significant (p = 0.004). Females had a significantly higher rate of anxiety (63.6%), and anxiety was more common in unmarried individuals (95.1%), those from families with two to five members (78.2%), and those with a family income between BDT 30,000 and BDT 60,000 (44.0%). Hostel residents (70.2%) and first-year medical students (24.9%) were also more likely to experience anxiety, although these differences were not statistically significant. The study found that stress was higher among female participants, unmarried individuals, those living in hostels, and those in the age group of 21 to 24 years and with a family size of two to five members. However, these differences were not statistically significant. The academic year of the participants was a significant predictor of stress, with first and second-year students reporting more stress than other years. Depression and anxiety were more prevalent in participants who did not have a habit of physical exercise, slept less than 8 hours, and were non-smokers. However, only physical exercise habit was significantly associated with depression and anxiety. Stress was also higher in participants without a habit of exercising, sleeping less than 8 hours, and not smoking, but the differences were not statistically significant.

**Table 2:** Association of depression, anxiety and stress with factors of the study participants.

| Variable | Depression | | $\chi^2$ | P-value | Anxiety | | $\chi^2$ | P-value | Stress | | $\chi^2$ | P-value |
| --- | --- | --- | --- | --- | --- | --- | --- | --- | --- | --- | --- | --- |
|  | Normal | Depressive symptoms |  |  | Normal | Anxiety symptoms |  |  | Normal | Stress symptoms |  |  |
| Socio-demographic factors |  |  |  |  |  |  |  |  |  |  |  |  |
| Age |  |  |  |  |  |  |  |  |  |  |  |  |
| ≤ 20 | 34<br>(23.1%) | 76<br>(35.9%) | 6.740<br>(df=2) | 0.034* | 27<br>(20.1%) | 83<br>(36.9%) | 11.074<br>(df=2) | 0.004* | 57<br>(27.0%) | 53<br>(35.8%) | 3.303<br>(df=2) | 0.192 |
| 21 – 24 | 106<br>(72.1%) | 129<br>(60.8%) |  |  | 101<br>(75.4%) | 134<br>(59.6%) |  |  | 146<br>(69.2%) | 89<br>(60.1%) |  |  |
| ≥ 25 | 7<br>(4.8%) | 7<br>(3.3%) |  |  | 6<br>(4.5%) | 8<br>(3.5%) |  |  | 8<br>(3.8%) | 6<br>(4.1%) |  |  |
| Gender |  |  |  |  |  |  |  |  |  |  |  |  |
| Male | 69<br>(46.9%) | 79<br>(37.3%) | 3.353<br>(df=1) | 0.067 | 66<br>(49.3%) | 82<br>(36.4%) | 5.687<br>(df=1) | 0.017* | 95<br>(45.0%) | 53<br>(35.8%) | 3.047<br>(df=1) | 0.081 |
| Female | 78<br>(53.1%) | 133<br>(62.7%) |  |  | 68<br>(50.7%) | 143<br>(63.6%) |  |  | 116<br>(55.0%) | 95<br>(64.2%) |  |  |
| Marital status |  |  |  |  |  |  |  |  |  |  |  |  |
| Unmarried | 138<br>(93.9%) | 200<br>(94.3%) | 4.231 | 0.105 | 124<br>(92.5%) | 214<br>(95.1%) | 1.711 | 0.460 | 199<br>(94.3%) | 139<br>(93.9%) | 0.306 | 0.918 |
| Married | 6<br>(4.1%) | 12<br>(5.7%) |  |  | 8<br>(6.0%) | 10<br>(4.4%) |  |  | 10<br>(4.7%) | 8<br>(5.4%) |  |  |
| Divorced/<br>Separated | 3<br>(2.0%) | 0<br>(0.0%) |  |  | 2<br>(1.5%) | 1<br>(0.5%) |  |  | 2<br>(1.0%) | 1<br>(0.7%) |  |  |
| Family size |  |  |  |  |  |  |  |  |  |  |  |  |
| Two to five<br>members | 118<br>(80.3%) | 168<br>(79.2%) | 2.708 | 0.244 | 110<br>(82.1%) | 176<br>(78.2%) | 1.305 | 0.526 | 166<br>(78.7%) | 120<br>(81.1%) | 0.686 | 0.801 |
| Six to ten<br>members | 27<br>(18.4%) | 44<br>(20.8%) |  |  | 23<br>(17.2%) | 48<br>(21.3%) |  |  | 44<br>(20.9%) | 27<br>(18.2%) |  |  |
| More than ten<br>members | 2<br>(1.3%) | 0<br>(0.0%) |  |  | 1<br>(0.7%) | 1<br>(0.5%) |  |  | 1<br>(0.4%) | 1<br>(0.7%) |  |  |
| Family income |  |  |  |  |  |  |  |  |  |  |  |  |
| Less than<br>BDT 30,000 | 30<br>(20.4%) | 47<br>(22.2%) | 0.997<br>(df=2) | 0.607 | 25<br>(18.7%) | 52<br>(23.1%) | 1.736<br>(df=2) | 0.420 | 44<br>(20.9%) | 33<br>(22.3%) | 0.376<br>(df=2) | 0.829 |
| Between BDT<br>30,000 and<br>BDT 60,000 | 73<br>(49.7%) | 94<br>(44.3%) |  |  | 68<br>(50.7%) | 99<br>(44.0%) |  |  | 101<br>(47.9%) | 66<br>(44.6%) |  |  |
| More than<br>BDT 60,000 | 44<br>(29.9%) | 71<br>(33.5%) |  |  | 41<br>(30.6%) | 74<br>(32.9%) |  |  | 66<br>(31.2%) | 49<br>(33.1%) |  |  |
| Residence |  |  |  |  |  |  |  |  |  |  |  |  |
| Hostel | 112<br>(76.2%) | 145<br>(68.4%) | 2.990<br>(df=2) | 0.224 | 99<br>(73.9%) | 158<br>(70.2%) | 0.963<br>(df=2) | 0.618 | 154<br>(73.0%) | 103<br>(69.6%) | 0.565<br>(df=2) | 0.754 |
| Home with<br>parents | 28<br>(19.0%) | 57<br>(26.9%) |  |  | 28<br>(20.9%) | 57<br>(25.3%) |  |  | 48<br>(22.7%) | 37<br>(25.0%) |  |  |
| Flat or<br>apartment | 7<br>(4.8%) | 10<br>(4.7%) |  |  | 7<br>(5.2%) | 10<br>(4.5%) |  |  | 9<br>(4.3%) | 8<br>(5.4%) |  |  |
| Academic year |  |  |  |  |  |  |  |  |  |  |  |  |
| First year | 24<br>(16.3%) | 53<br>(25.0%) | 9.200<br>(df=4) | 0.056 | 21<br>(15.7%) | 56<br>(24.9%) | 9.362<br>(df=4) | 0.053 | 44<br>(20.8%) | 33<br>(22.3%) | 12.053<br>(df=4) | 0.017* |
| Second year | 23<br>(15.7%) | 48<br>(22.6%) |  |  | 21<br>(15.7%) | 50<br>(22.2%) |  |  | 30<br>(14.2%) | 41<br>(27.7%) |  |  |
| Third year | 34<br>(23.1%) | 34<br>(16.0%) |  |  | 32<br>(23.9%) | 36<br>(16.0%) |  |  | 42<br>(19.0%) | 26<br>(17.6%) |  |  |
| Fourth year | 30<br>(20.4%) | 37<br>(17.5%) |  |  | 27<br>(20.1%) | 40<br>(17.8%) |  |  | 46<br>(21.8%) | 21<br>(14.2%) |  |  |
| Fifth year | 36<br>(24.5%) | 40<br>(18.9%) |  |  | 33<br>(24.6%) | 43<br>(19.1%) |  |  | 49<br>(23.2%) | 27<br>(18.2%) |  |  |
| **Behavioral factors** |  |  |  |  |  |  |  |  |  |  |  |  |
| Physical exercise |  |  |  |  |  |  |  |  |  |  |  |  |
| No | 82<br>(55.8%) | 153<br>(72.1%) | 10.311<br>(df=1) | **0.001\*** | 78<br>(58.2%) | 157<br>(69.8%) | 4.971<br>(df=1) | **0.026\*** | 133<br>(63.0%) | 102<br>(68.9%) | 1.333<br>(df=1) | 0.248 |
| Yes | 65<br>(44.2%) | 59<br>(27.9%) |  |  | 56<br>(41.8%) | 68<br>(30.2%) |  |  | 78<br>(37.0%) | 46<br>(31.1%) |  |  |
| Hours of sleep |  |  |  |  |  |  |  |  |  |  |  |  |
| Less than 8 hours | 112<br>(76.2%) | 159<br>(75.0%) | 0.066<br>(df=1) | 0.797 | 104<br>(77.6%) | 167<br>(74.2%) | 0.522<br>(df=1) | 0.470 | 159<br>(75.4%) | 112<br>(75.7%) | 0.005<br>(df=1) | 0.945 |
| More than 8 hours | 35<br>(23.8%) | 53<br>(25.0%) |  |  | 30<br>(22.4%) | 58<br>(25.8%) |  |  | 52<br>(24.6%) | 36<br>(24.3%) |  |  |
| Smoking status |  |  |  |  |  |  |  |  |  |  |  |  |
| Not smoking | 140<br>(95.2%) | 208<br>(98.1%) |  | 0.132 | 127<br>(94.8%) | 221<br>(98.2%) |  | 0.109 | 202<br>(95.7%) | 146<br>(98.6%) |  | 0.133 |
| Smoking | 7<br>(4.8%) | 4<br>(1.9%) |  |  | 7<br>(5.2%) | 4<br>(1.8%) |  |  | 9<br>(4.3%) | 2<br>(1.4%) |  |  |
| **COVID-19 related factors** |  |  |  |  |  |  |  |  |  |  |  |  |
| Previous COVID-19 infection status |  |  |  |  |  |  |  |  |  |  |  |  |
| Not infected before | 108<br>(73.5%) | 151<br>(71.2%) | 0.217<br>(df=1) | 0.641 | 104<br>(77.6%) | 155<br>(68.9%) | 3.180<br>(df=1) | 0.075 | 159<br>(75.4%) | 100<br>(67.6%) | 2.625<br>(df=1) | 0.105 |
| Infected before | 39<br>(26.5%) | 61<br>(28.8%) |  |  | 30<br>(22.4%) | 70<br>(31.1%) |  |  | 52<br>(24.6%) | 48<br>(32.4%) |  |  |
| Vaccination status |  |  |  |  |  |  |  |  |  |  |  |  |
| Unvaccinated | 2<br>(1.4%) | 2<br>(0.9%) | 1.051 | 0.760 | 0<br>(0.0%) | 4<br>(17.8%) | 2.138 | 0.414 | 2<br>(1.0%) | 2<br>(1.4%) | 0.379 | 1.000 |
| Vaccinated (single dose) | 1<br>(0.7%) | 4<br>(1.9%) |  |  | 2<br>(1.5%) | 3<br>(13.3%) |  |  | 3<br>(1.4%) | 2<br>(1.4%) |  |  |
| Vaccinated (both dose) | 144<br>(97.9%) | 206<br>(97.2%) |  |  | 132<br>(98.5%) | 218<br>(96.9%) |  |  | 206<br>(97.6%) | 144<br>(97.2%) |  |  |
| Total | 147<br>(100%) | 212<br>(100%) |  |  | 134<br>(100%) | 225<br>(100%) |  |  | 211<br>(100%) | 148<br>(100%) |  |  |

\* Pearson chi-squared test was done.

Although participants who were not previously infected with COVID-19 and had received both doses of the vaccine showed comparatively more depression, anxiety, and stress, the COVID-19 related factors did not show any significant association with these mental health issues.

## Discussion

Medical students have always been subjected to more psychological abuse than others, given that medical science has one of the most difficult course curriculums [5]. Mental health has been under question of everyone with the rise of COVID-19 pandemic. Medical students with their prevalent poor mental health before COVID-19, is under dilemma about their mental health amid the COVID-19 pandemic.

This study’s findings revealed an alarmingly high prevalence of poor mental health, which is comparable to a study conducted in 12 nations [8]. This shows significant rise of poor mental health during the COVID-19 pandemic when compared to previous study by Hasan et al [13]. Anxiety came in the most prevalent mental health disorder, followed by depression in this study participants. Around 3 out of 5 medical students had either depression or anxiety, which is consistent with other studies [14], while 2 out of 5 medical students had stress.

Given the current pandemic, financial crisis and educational burden, such a high proportion of poor mental health is to be foreseen. In this study, female students were shown to be more likely to suffer from depression, anxiety, and stress, with anxiety having the significant link. This is consistent with other studies [10,15–17] but contradicts the study by Hasan et al [13]. where male predominance was seen for poor mental health. Younger medical students were associated with depressive along with anxiety symptoms. Other study conducted in United Kingdom[10] had similar findings with younger medical students and junior doctors having poorer mental health when compared to seniors.

New information keeps on adding with elevating medical academic year but joining the medical studies for first time has feeling of whole new world. New environment, new friends, new teacher and different level of educational expectation affects the mental health of medical students in their freshman years. Increase in academic year showed relatively decrease in depression, anxiety and stress in this study. This is easily expected as medical students in first year become used to with the medical educational system in the later years.

Other studies also show similar findings regarding the academic year of medical students and their mental health [13,18,19]. Interestingly, the during COVID-19 pandemic, doing physical exercise was linked to decreased depression and anxiety. Other studies among medical students support this [20,21].

In conclusion, poor mental health is still highly prevalent in the medical students.

Depression, anxiety and stress has always been terrorizing the medical students and COVID-19 pandemic has kept adding the more misery. Different factors like age, gender, academic year, and physical exercise behavior have affected medical students’ mental health. This information is very crucial in order to create a welcoming environment for medical students. Attention should be given towards the needs of the more vulnerable demographics like the female students and the medical students in first and second year. Screening and counselling program should be conducted regularly in timely interval. Self-care is very essential to preserve a good mental health. Further study is essential to see the bigger picture of the mental health of the medical students as this study had big time constraint.

## Data Availability

All relevant data are within the manuscript.

## Acknowledgement

We would like to express our gratitude to Monidipa Saha, our supervisor, for making this study possible. Her expertise and assistance guided us through every step of the process of planning and completing our capstone research project. We would also want to convey our gratitude to all of the medical students who participated in this study despite the tough circumstances of the COVID-19 outbreak. We would also like to give special thanks to Dr. Tariqul Islam, Intern doctor, Dhaka Medical College Hospital for his valuable time and kind support during data collection phase. He helped us a lot to get the proper timing and class schedule for the data collection.

